# A package of interventions to increase facility births attended by a birth companion in Ethiopia, Kenya, and Nigeria: A cluster-randomized controlled trial

**DOI:** 10.1101/2025.08.07.25333264

**Authors:** Gadise Bekele Regassa, Patricia Owira, Melanie Olum, Felagot Taddese, Konjit Wolde, Osborn Koech Kiptoo, Lisa Noguchi, Anne Hyre, Samuel Mwaura, Dedefo Teno, Walelegn Worku, Alemayehu Worku, Jefferson Mwaisaka, Abel Mokua, Stephanie Suhowatsky, Delayehu Bekele, Solomon Getachew Alem, Pooja Sripad, Gayane Yenokyan, Oluwatosin Laleye, Mobolanle Balogun, Zubaida Farouk, Adeola Folashade Afolabi, Eberechukwu Eke, Jennyfer Don Aki, Della Berhanu

## Abstract

**Background:** Despite the World Health Organization’s recommendation of offering women the option to have a birth companion of their choice, implementation of this recommendation remains limited in low- and middle-income countries, including Ethiopia, Kenya, and Nigeria. This study assessed if a tailored set of programmatic interventions targeting common barriers to birth companionship could increase the proportion of women giving birth with a companion.

**Methods:** In this cluster-randomized trial,12 facilities in Ethiopia, 24 facilities across Machakos and Murang’a counties in Kenya, and 24 facilities in Kano and Nasarawa states in Nigeria were randomized (3:1) to intervention and control arms (ClinicalTrials.gov NCT05565196). The intervention was introduced over a period of two months and was evaluated after 12 additional months of implementation. We enrolled women having vaginal births. Data were collected at baseline, quarterly, and one year, using a facility survey and qualitative interviews with women, birth companions, health workers, and unit managers. We used a cluster-weighted chi-square test to compare intervention and control arms with respect to the primary outcome, which was the proportion of women accompanied in the facility by a companion during labor, birth, and postnatal periods. Difference-in-difference analysis was also done.

**Findings:** 1,475 women in Ethiopia, 1,496 in Machakos and 1,521 in Murang’a Counties of Kenya and 1,644 in Kano and 1,249 in Nasarawa states of Nigeria were recruited. At end-line, primary outcome increased in intervention arm by 38% points (95%CI=12-66, p=0.009) in Ethiopia,15% points (95%CI=7-26, p=0.002) in Murang’a, 9% points in Machakos (95%CI=-1-20, p=0.077), 51%(95%CI=9-93, p=0.018) points in Kano state, and 53% points in Nasarawa state (95%CI=11-96, 0.014).

**Interpretation:** A tailored interventions reduced barriers to women having continuous companionship in facilities during labor, birth, and the postnatal period and has potential for use in a range of facility types and settings aiming to increase client-centered intrapartum care.

## Introduction

Maternal and neonatal morbidity and mortality remain unacceptably high in many parts of the world, [1,2] underscoring the need for targeted interventions to improve maternal and newborn health (MNH) outcomes. The presence of a birth companion during a woman’s labor and birth represents a simple and low-cost strategy that can improve MNH outcomes [3].

The World Health Organization (WHO) defines a birth companion as any person chosen by the woman to provide her with continuous support during labor and childbirth [4]. Support can include: offering emotional support through continuous presence; giving praise and reassurance; offering information updates on the progress of labor; providing coping strategies such as proper breathing techniques; comforting through massage; aiding in mobility; reminding the woman to drink fluids and use the restroom; advocating for her needs; and participating in newborn care [3,5,6].

Research in different geographies has consistently demonstrated that women greatly value and derive significant benefits from having a trusted support person present throughout labor and birth [6]. Women who have a companion of choice during labor and birth report this support helps them feel safe, strong, confident, and secure [7]. Women with continuous support during labor and birth are more likely to have shorter labor, spontaneous vaginal births, improved newborn APGAR scores, and increased satisfaction with the care they receive. They are also less likely to need pain medication during labor or have caesarean deliveries [3,5,6]. As such, the WHO recommends “continuous companionship during labor and childbirth” [8] and advises that all women have a companion of their choice throughout labor and childbirth [9].

WHO recommendations [4,9] have not led to birth companionship as routine practice in most sub-Saharan African countries [10–14]. Countries with supportive national policies and guidelines lack widespread adoption of birth companionship [14,15]. For example, Ministries of Health in Ethiopia and Kenya have national guidelines that support birth companionship and prioritize quality of care [16,17]. Both countries report a substantial rise in facility births (in Ethiopia from 10% in 2011 to 48% in 2019; in Kenya 48% in 2008 to 89% in 2022), mainly occurring in public health facilities [16–19], yet birth companionship is not prevalent. A study in Ethiopia reported that 98% of women were not allowed to have a birth companion [12], while in Kenya another study reported that 78% of women were not permitted to have a companion during labor, and 84% were denied companionship during birth [13]. In Nigeria, where delivery by a skilled attendant has a relatively slower increase (from 37% in 2003 to 46% in 2024)[20] there is no explicit inclusion of birth companions in national clinical guidelines, however, birth companions are included in the 2015 Respectful Maternity Care Toolkit and Trainers’ Guide[21]. A study in Nigeria reported that 29% of women reported having a companion during labor, childbirth and after childbirth [22]. Several barriers contribute to the limited implementation of birth companionship within routine practice. These barriers include restrictive health facility policies, negative attitudes of healthcare providers, inadequate facility infrastructure, and insufficient preparation for families and birth companions [12,23].

The paucity of evidence on effective implementation strategies to address key barriers to birth companionship [24,25] in facilities suggests further research is needed to design and evaluate models that can overcome known barriers. These models should consider local context and the values and preferences of both women and providers [7] to promote greater uptake of birth companions in health centers and hospitals across diverse clinical settings and contexts [24]. To the best of our knowledge, not many studies have directly assessed the effect of implementing a tailored programmatic intervention on coverage of facility births attended by a companion. The intervention package that we tested in this trial was based on the logic model described in a Cochrane review that synthesized qualitative evidence and reviewed interventions relevant to continuous support to women during childbirth [7]. Here, we aimed to evaluate the impact of a tailored intervention package on the proportion of facility births with a companion present in the labor, birth, and postnatal periods.

## METHODS

### Design and setting

This study was a two-parallel arm, cluster-randomized controlled trial (cRCT) that included 11 public health facilities in Addis Ababa and one in Oromia, Ethiopia; 24 public health facilities in rural and semi-rural Machakos and Murang’a counties of Kenya; and, 24 health facilities in Kano and Nasarawa states in Nigeria. In Ethiopia, all facilities were health centers and 11 were located in the capital city and one in the outskirts of the city, in Oromia. All facilities were served by a tertiary referral facility, St. Paul’s Hospital, which had integrated birth companions into routine care. In Kenya and Nigeria, both health centers and hospitals were included.

### Ethics approval

This study was approved by the Addis Continental Institute of Public Health Institutional Review Board (IRB) (FWA 00015151) in Ethiopia, Amref Health Ethical Review Committee (FWA 00018713) in Kenya, the National Health Research Ethics Committee (NHREC/01/01/2007) in Nigeria, and the John Hopkins Bloomberg School of Public Health IRB (No 21183) in the United States. All methods were carried out in accordance with relevant guidelines and regulations of the study countries. Verbal informed consent from participants for both quantitative and qualitative components was taken and witnessed and recorded by the data collector.

Confidentiality was kept to the maximum standard during data collection and data handling. The study protocol has been published [26] and the study is registered with ClinicalTrials.gov (NCT05565196), first posted on October 4, 2022.

### Facilities

In Ethiopia, facilities were selected in consultation with the Addis Ababa Regional Health Bureau and St. Paul’s Hospital, in Kenya in consultation with the respective County Health Management Teams, and in Nigeria, in consultation with the respective State Ministry of Health and State Primary Healthcare Development Agency. Facility eligibility criteria are detailed in the protocol paper [26]. Briefly, facilities were classified as low- (<23 births/month in Ethiopia, <20 births/month in Kenya, <50 births/month in Nigeria), medium- (23-84 births/month in Ethiopia, 20-60 births/month in Kenya, 50-179 births/month in Nigeria), or high-volume (>84 births/month in Ethiopia, >60 births/month in Kenya, >179 births/month in Nigeria). In Ethiopia, there were 12 facilities within the catchment area of St. Paul’s Hospital, which were all included in the study. In Kenya, 36 facilities in Machakos and 20 in Murang’a were eligible. In Machakos county, a high-volume facility was replaced before randomization due to the management’s concern about feasibility of the intervention given high client load and limited space. In Nigeria, 27 facilities in Kano state and 19 facilities in Nasarawa state were eligible. Within each category, the four facilities with the highest deliveries were identified.

### Participants

Eligibility criteria for participants are detailed in the published protocol [26]. Briefly, for the quantitative survey, eligible women aged 15 years or older who gave birth vaginally at participating facilities, could provide informed consent and were discharged during the day were included. In Nigeria, women discharged at night were also included. For the qualitative component, eligible participants included women, birth companions, healthcare providers in antenatal, labor, birth, and postnatal wards, and unit managers present during the study. Participants self-reported sex as male or female.

Women giving birth in study facilities were referred by providers to study investigators, who assessed eligibility and obtained consent before or after birth. Some women in intervention facilities participated in quarterly in-depth interviews (IDIs), including those reporting having and not having positive experiences of birth in the quantitative interviews. Birth companions were recruited, including husbands, other family members, and friends. Facility directors identified providers for quarterly focus group discussions (FGDs). Unit managers from the antenatal, labor, and birth wards were also invited for quarterly key informant interviews (KIIs). We approached health system managers overseeing intervention facilities for endline KII.

### Randomization and masking

Johns Hopkins Biostatistics Center conducted the randomization of facilities after a four-week baseline assessment. Facilities were randomized to the intervention or control arm in a 3:1 ratio, with stratification by the delivery volume categories, country, additionally in Kenya by county and in Nigeria by state. The randomization was weighted in favor of the intervention due to existing evidence of benefit for birth companionship. We implemented the intervention package over two months. Control facilities continued to provide the local standard of care. No masking of the intervention occurred in this study because the nature of the intervention required active involvement from both participants and providers.

### Procedures

This study evaluated an intervention package of eight elements to increase birth companion coverage, based on the identified barriers in the Cochrane review [7]. Components of the intervention are shown in Table 1.

**Table 1:**
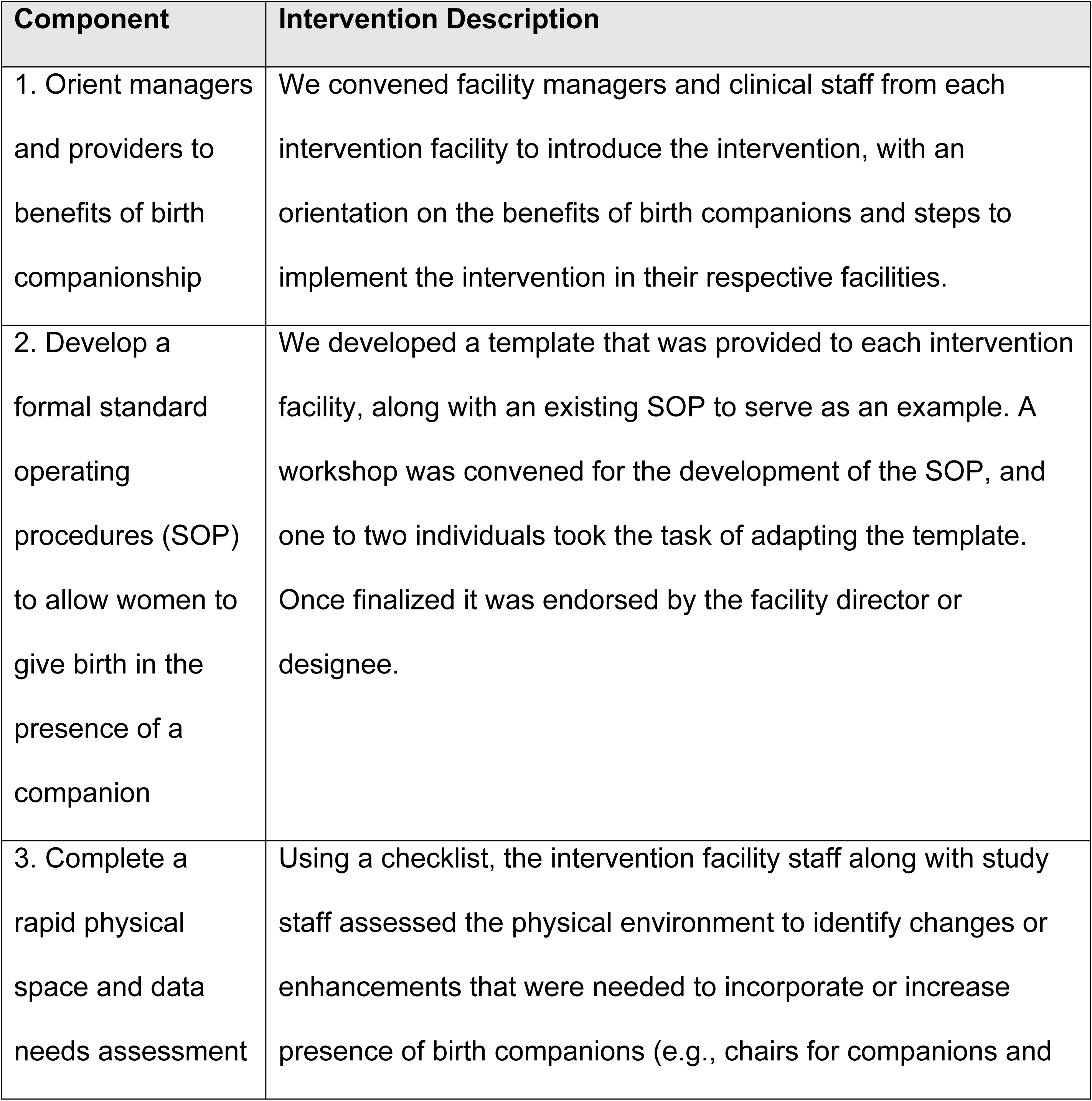
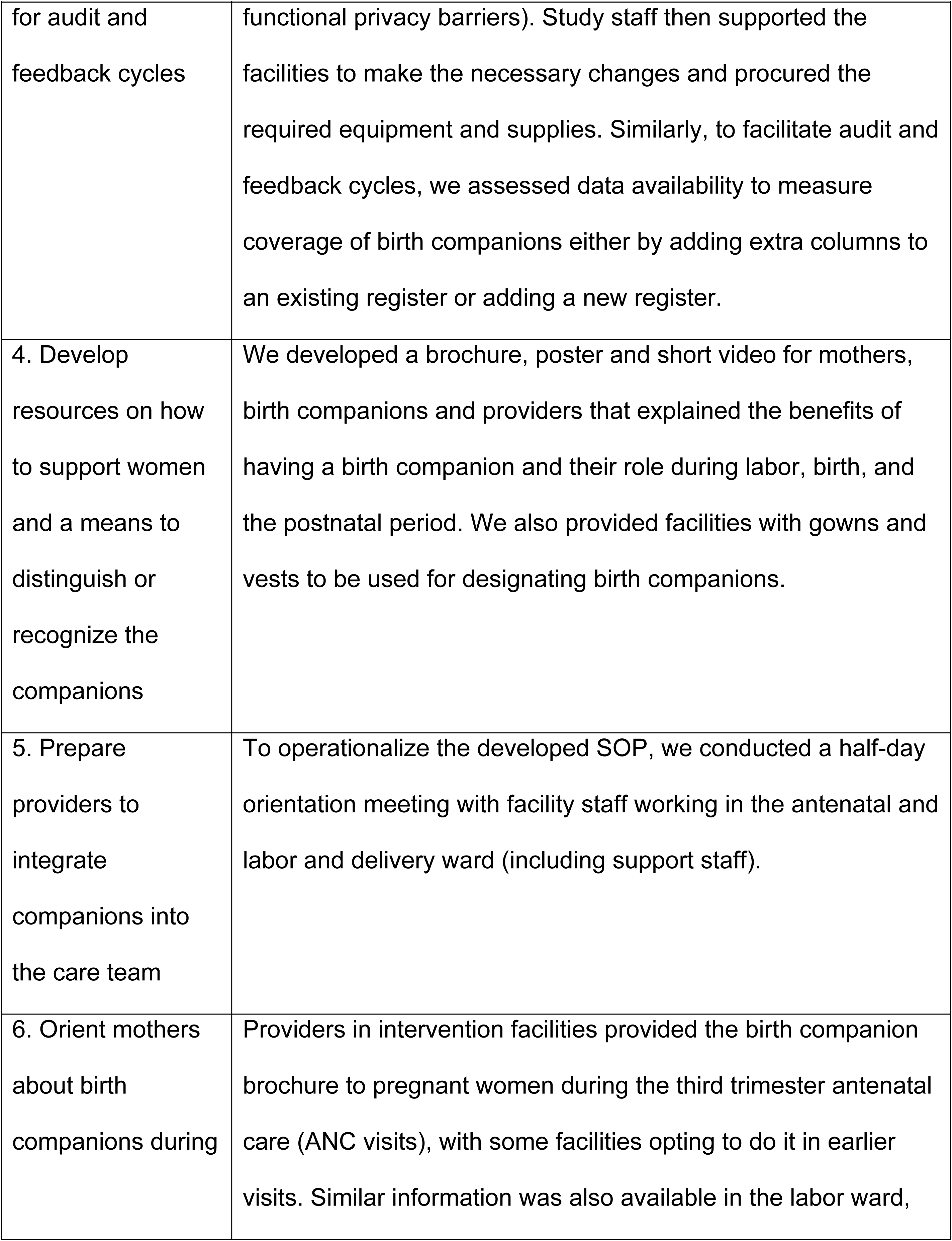
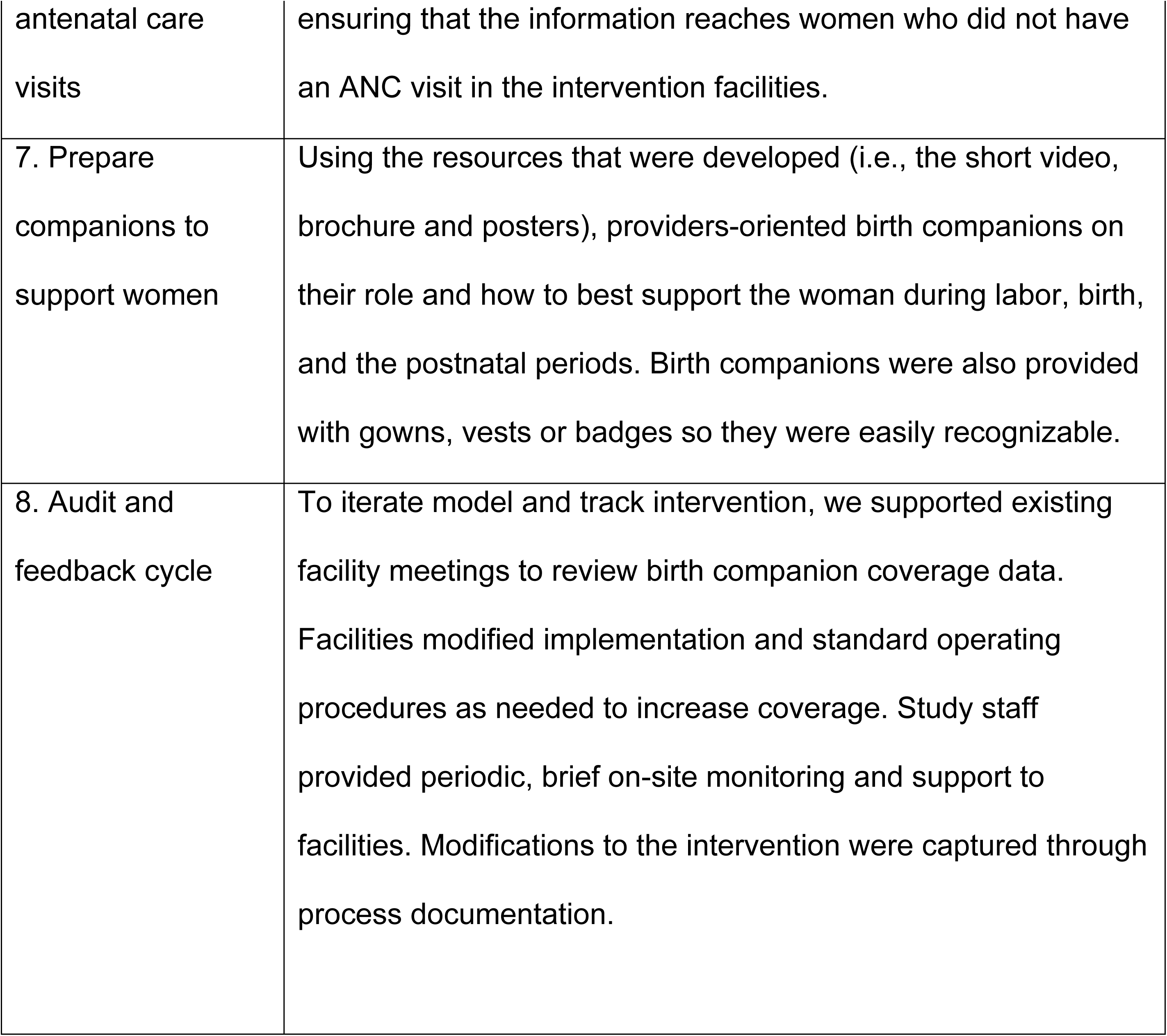
Components of the birth companion intervention package

Study participation was voluntary. We did not use retention strategies in the study, as assessment of the primary outcome occurred cross-sectionally for births occurring at five timepoints during study implementation. Study staff provided on-site monitoring and support to implement the intervention package effectively. Monitoring and support visits lasted one to two hours per facility and the visits were more frequent at the start of the intervention (every two weeks in Ethiopia and monthly in Kenya) and decreased to once a quarter after six months of implementation. In Nigeria monitoring and support visits occurred monthly in the first two months of implementation then reduced to every two months and quarterly as the intervention progressed.

### Data collection

#### Quantitative

Baseline data collection was conducted in 2022, from October 10 to November 6 in Ethiopia, from November 1 to December 9 in Kenya, and from August 14 to September 12 in Nigeria. Following a two-month period to introduce the intervention, we conducted three quarterly surveys, each lasting one week. In 2024, we conducted a final survey from February 5 to March 3 in Ethiopia, March 19 to June 14 in Kenya, and October 21 to November 18 in Nigeria. Data collection in Kenya was paused in April due to a health providers’ strike that lasted nearly two months.

#### Qualitative

Following the baseline survey, three rounds of qualitative data collection were conducted quarterly in intervention facilities. In Ethiopia, an additional fourth round of qualitative data was conducted at endline.

### Outcomes

#### Primary outcome

The primary outcome was defined as the proportion of women at study facilities that had a companion present during labor, birth, and the postnatal (continuously at all timepoints) period. It was assessed separately for Ethiopia, Kenya, and Nigeria.

#### Secondary outcomes

Among those who reported having a birth companion, we measured the proportion reporting that the birth companion was the one of their choices. We assessed experience of care among birthing women using the Person-Centered Maternity Care (PCMC) [27] scale and qualitatively through women’s, birth companions’, providers’ and unit managers’ perception of birth companionship.

### Statistical analysis and power

#### Sample size

The sample size aimed to estimate birth companionship coverage using an intra-class correlation coefficient (ICC) between 0.05 and 0.5. With post-birth companion support assumed at 50% and baseline coverage between 5% and 20%, the sample size was calculated for the three countries using PASS 15 Power Analysis and Sample Size Software (2017) [28]. To achieve 80% statistical power at a 0.05 significance level, assuming 20% coverage in the control arm and 50% in the intervention arm, 62 women per facility were needed, totaling approximately 744 women across both arms. The sample size accounted for varying cluster sizes.

Qualitative data were collected through purposive sampling. In each country, quarterly IDIs were conducted with 16 women and 16 birth companions in intervention facilities to assess experiences and the intervention’s acceptability. Women who had low and high PCMC scores were included. Six provider FGDs and six KIIs with facility managers were held quarterly in Ethiopia and Kenya, while eight provider FGDs and eight KIIs with unit managers were held quarterly in Nigeria to explore acceptability and feasibility of birth companionship. In Ethiopia only, nine KIIs with health system managers were conducted at endline to assess the intervention’s sustainability.

### Analysis

To characterize and compare intervention and control arms, we calculated proportions for categorical variables and mean and standard deviation for continuous variables.

#### Analysis of primary outcomes

The primary outcome was compared between intervention and control arms at the end of the intervention using a cluster-weighted chi-square test. We adjusted for covariates that were different between intervention and control clusters at endline using a logistic regression model with cluster-correlated standard error estimates to account for within-facility correlation of outcomes. To account for differences in the primary outcome at baseline, we also conducted a difference-in-difference analysis using a linear regression, adjusting for clustering and factors that were different in intervention and control clusters at baseline and endline. The regression included an interaction term for the timing of the survey (baseline or endline) and the facility allocation (intervention or control cluster), measuring the intervention’s effect as a percentage point difference with 95% confidence intervals (95% CI). Additionally, we assessed graphically the trends in proportion of births meeting the primary outcome in intervention and control facilities over the five data collection periods.

#### Analysis of secondary outcomes

For the secondary outcome of having a birth companion of choice, we compared women’s preferred companion to the one reported. We measured experience of care using the PCMC scale normalized to 100 before analysis and compared mean scores in intervention and control clusters using a cluster-weighted t-test. Linear regression models with cluster-correlated standard error estimates were used to compare intervention and control arms while adjusting for covariates that were different between them at endline. The validated 33-item PCMC scale included questions about birth companionship, some of which overlapped with those used to measure our primary outcome. To avoid redundancy, we removed these questions.

Recognizing that this adjustment might impact the tool’s validity, we report on the validated 13-item PCMC scale,[29] which measures women’s experience of care without including questions about birth companions. Stata 15 was used for analyses[30].

#### Qualitative analysis

Audio recordings of IDI and FGD were transcribed from local languages to English by individuals proficient in these languages. Thematic analyses were operationalized by application of an inductively and deductively informed coding structure to textual passages using Atlas.ti 7.5 [31] and 24.0 [32] for Ethiopia and Nigeria respectively, and Dedoose 9.0.17 [33] for Kenya. In Ethiopia, GBR and SGA, in Kenya, MO and JM independently, and in Nigeria OL reviewed all transcripts and field notes. They identified themes that were then aligned with research questions on experience of care. Data analysis followed a pattern of association on emerging themes focusing on participants’ perceptions. Additional qualitative data will be presented in a planned, separate manuscript.

## RESULTS

Recruitment occurred between October 10, 2022 and November 18, 2024. In Ethiopia, a total of 1,457 women were enrolled from 12 facilities (clusters) in the Addis Ababa and Oromia regions (Fig 1). In the three quarterly surveys, each lasting one week, we included: 147, 130, and 170 women in intervention facilities; and 40, 36, and 29 women in control facilities. At baseline and endline in Kenya, we enrolled 3,017 women across 24 facilities in Machkso and Muran’a counties (Fig 1). All screened women were enrolled in Kenya study. Across three quarterly surveys, in Machakos we included: 149, 137, and 131 women in intervention facilities; and 36, 51, and 39 women in control facilities. Similarly, in Murang’a we included 130, 135, and 118 women in intervention and 60, 66, and 42 in women in control facilities. In Nigeria, a total we of 2,864 women were enrolled over the study period form 24 facilities in Kano and Nasarawa states (Fig 1). Across the quarterly surveys, in Kano, we included: 141, 147, and 171 women in intervention facilities and 79, 67.and 66 women in control facilities. Similarly, in Nasarawa, we included: 94, 163, and 113 women in intervention facilities, and 34, 31, and 23 women in control facilities. In Nigeria, data was not collected at baseline from one of the intervention facilities in Kano state due to an error in locating the correct facility; and at endline, we could not collect data from one of the control facilities because it was under renovation throughout the data collection period (Fig 1).

**Fig 1:**
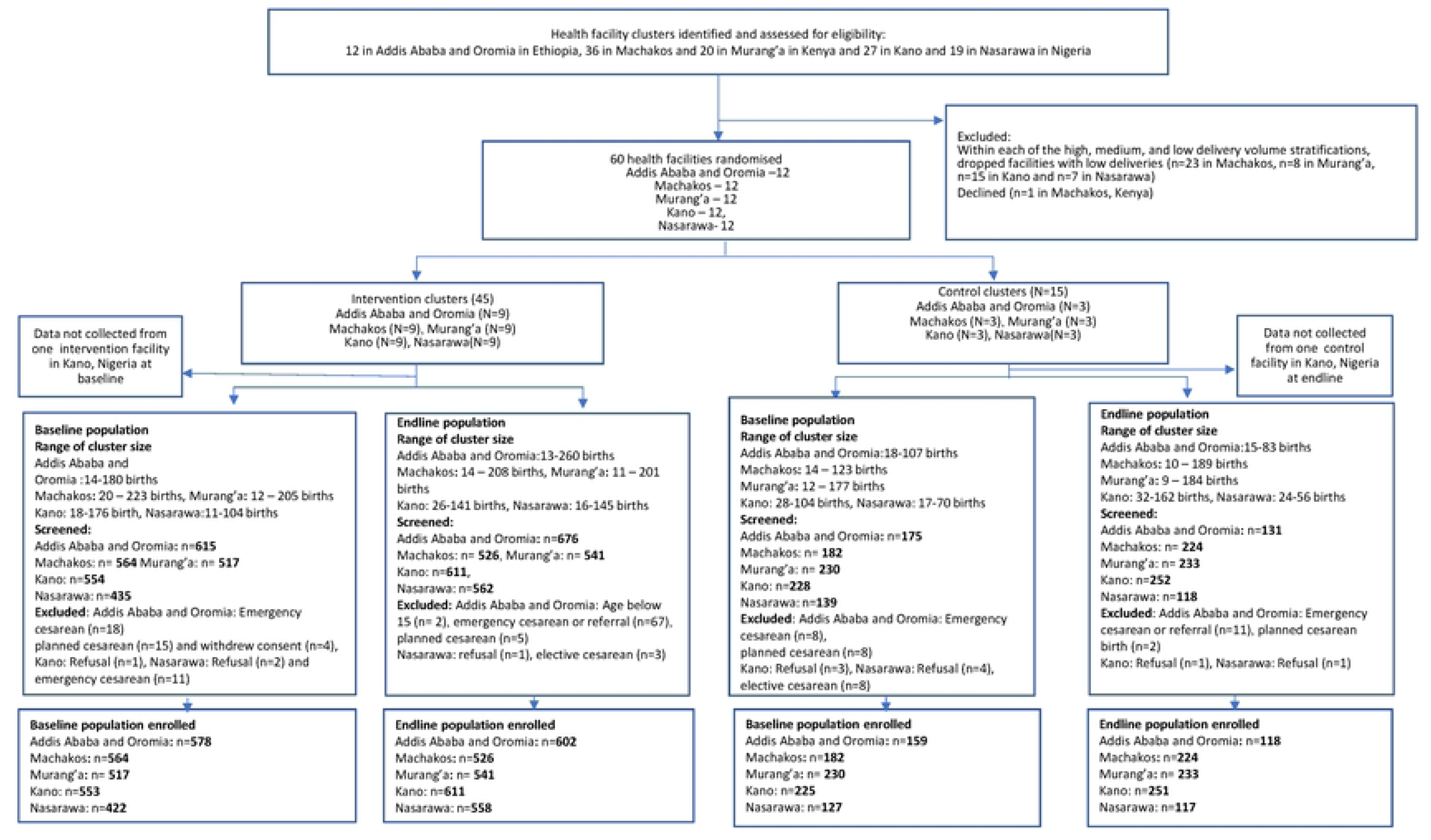
Trial Profile in Ethiopia, Kenya, and Nigeria

In Ethiopia, using cutoff points based on average monthly deliveries prior to the baseline survey, resulted in a 3:1 ratio of intervention and control facilities in each of the low, medium and high delivery categories, whereas in in Kenya and Nigeria fewer facilities fell in the low delivery volume category. Relatively equal number of facilities offered comprehensive emergency obstetric and newborn care (CEmONC), and basic emergency obstetric and newborn care (BEmONC).

In Ethiopia, the average age of women at baseline in both arms was 27 years. Over 90% of women were currently married and living with their partners. Half of women had a primary education. In both Machakos and Murang’a counties, seven out of ten women were married or cohabiting, and over 40% had a secondary education. In Nigeria women were similar in mean age, almost all women were married or living with their partners and most of the women had secondary education across intervention and control groups in both states (Table 2).

**Table 2:** Characteristics of study facilities and birthing women participating in the baseline survey in Ethiopia, Kenya and Nigeria

|  | Facilities |  |  |  |  |  |  |  |  |  |
| --- | --- | --- | --- | --- | --- | --- | --- | --- | --- | --- |
|  | Ethiopia |  | Kenya |  |  |  | Nigeria |  |  |  |
|  | Addis<br>Ababa/Oromia |  | Machakos |  | Murang'a |  | Kano |  | Nasarawa |  |
|  | Interven<br>tion<br>N=9 | Control<br>N=3 | Interven<br>tion<br>N=9 | Control<br>N=3 | Interv<br>ention<br>N=9 | Control<br>N=3 | Interven<br>tion N=9 | Cont<br>rol<br>N=3 | Interven<br>tion N=9 | Control<br>N=3 |
|  | n (%) | n (%) | n (%) | n (%) | n (%) | n (%) | n (%) | n (%) | n (%) | n (%) |
| **Birth volume** |  |  |  |  |  |  |  |  |  |  |
| Low<sup>a</sup> | 3 (33) | 1 (33) | 0 (0) | 1 (33) | 2 (22) | 1 (33) | 2 (22) | 0 (0) | 1 (33) | 1 (33) |
| Medium<sup>b</sup> | 3 (33) | 1 (33) | 6 (67) | 1 (33) | 3 (33) | 1 (33) | 2 (22) | 2<br>(67) | 3 (33) | 1 (33) |
| High<sup>c</sup> | 3 (33) | 1 (33) | 3 (33) | 1 (33) | 4 (44) | 1 (33) | 5 (56) | 1<br>(33) | 5 (56) | 1 (33) |
| **Type of**<br>**facility** |  |  |  |  |  |  |  |  |  |  |
| Health center | 9 (100) | 3(100) | 5 (56) | 1 (33) | 4 (44) | 1 (33) | 6 (67) | 2<br>(67) | 4 (44) | 1 (33) |
| Hospital | 0 | 0 | 4 (44) | 2 (67) | 5 (56) | 2 (67) | 3 (33) | 1<br>(33) | 5 (56) | 2 (67) |
| **Type of**<br>**services** |  |  |  |  |  |  |  |  |  |  |
| BEmONC<sup>d</sup> | 7(78) | 2(67) | 3 (33) | 1 (33) | 5 (56) | 2 (67) | 3 (33) | 1<br>(33) | 5 (56) | 2 (67) |
| CEmONC<sup>e</sup> | 2 (22) | 1 (33) | 6 (67) | 2 (67) | 4 (44) | 1 (33) | 6 (67) | 2<br>(67) | 4 (44) | 1 (33) |
|  | **Birthing women** |  |  |  |  |  |  |  |  |  |
|  | Ethiopia |  | Kenya |  |  |  | Nigeria |  |  |  |
|  | Addis<br>Ababa/Oromia |  | Machakos |  | Murang'a |  | Kano |  | Nasarawa |  |
|  | Interven<br>tion<br>N= 578 | Control<br>N=159 | Interven<br>tion<br>N=564 | Control<br>N=182 | Interv<br>ention<br>N=<br>517 | Control<br>N=230 | Interven<br>tion<br>N=553 | Cont<br>rol<br>N=22<br>5 | Interven<br>tion N=<br>422 | Control<br>N=127 |
|  | Mean (S<br>D) | Mean (SD) | Mean (SD) | Mean (SD) | Mean (SD) | Mean (SD) | Mean (SD) | Mean (SD) | Mean (SD) | Mean (SD) |
| **Age** | 27 ± 4•5 | 27 ± 4•2 | 26 ± 6•0 | 30 ± 6•1 | 28 ± 6•4 | 26 ± 6•5 | 27 ± 6•0 | 28 ± 6•0 | 28.9 ± 6•3 | 27.1 ± 5•5 |
|  | n (%) | n (%) | n (%) | n (%) | n (%) | n (%) | n (%) | n (%) | n (%) | n (%) |
| **Marital status<sup>f</sup>** |  |  |  |  |  |  |  |  |  |  |
| Single | 6 (1) | 1 (1) | 136 (24) | 46 (25) | 96 (19) | 45 (20) | 0 (0) | 0 (0) | 5 (1.2) | 2 (1.6) |
| Married/Cohabiting | 565 (98) | 158 (99) | 419 (74) | 132 (73) | 377 (73) | 169 (74) | 547 (99) | 225 (100) | 417 (99) | 123 (97) |
| Not together<sup>g</sup>/Widowed | 7 (1) | 0 (0) | 8 (1) | 4 (2) | 8 (2) | 5 (2) | 4 (0.7) | 0 (0) | 0 (0) | 1 (0.8) |
| **Education** |  |  |  |  |  |  |  |  |  |  |
| None | 41 (7) | 14 (9) | 0 (0) | 3 (2) | 3 (1) | 3 (1) | 0 (0) | 0 (0) | 0 (0) | 0 (0) |
| Primary | 302 (52) | 81 (51) | 159 (28) | 59 (32) | 153 (30) | 78 (34) | 99 (18) | 35 (16) | 59 (14) | 16 (13) |
| Secondary | 150 (26) | 48 (30) | 307 (55) | 81 (45) | 255 (49) | 125 (54) | 299 (54) | 143 (64) | 231 (55) | 63 (50) |
| Tertiary | 63 (11) | 15 (9) | 97 (17) | 39 (21) | 95 (18) | 24 (10) | 48 (8.7) | 15 (6.7) | 36 (8.5) | 18 (14) |
| Other | 22 (4) | 1 (1) | 0 (0) | 0 (0) | 11 (2) | 0 (0) | 107 (19) | 32 (14) | 96 (23) | 30 (24) |
| **Religion<sup>h</sup>** |  |  |  |  |  |  |  |  |  |  |
| Christian (Orthodox Catholic, Protestant) | 396 (68) | 69 (43) | 554 (98) | 179 (98) | 502 (97) | 222 (97) | 19 (3.4) | 0 (0) | 204 (48) | 53 (42) |
| Muslim | 180 (31) | 90 (57) | 6 (1) | 0 (0) | 5 (1) | 0 (0) | 534 (97) | 255 (100) | 218 (52) | 71 (56) |
| Other | 2 (<1) | 0 (0) | 3 (1) | 3 (2) | 10 (2) | 8 (3) | 0 (0) | 0 (0) | 0 (0) | 3 (2.4) |
| **Wealth quintile** |  |  |  |  |  |  |  |  |  |  |
| Q1 (Lowest) | 43 (8) | 25 (16) | 148 (26) | 47 (26) | 93 (18) | 46 (20) | 109 (20) | 68 (30) | 93 (22) | 19 (15) |
| Q2 | 128 (22) | 61 (38) | 100 (18) | 46 (25) | 91<br>(18) | 52 (23) | 75 (14) | 48<br>(21) | 88 (21) | 33 (26) |
| Q3 | 139 (24) | 21 (13) | 89 (16) | 31(17) | 92<br>(18) | 48 (21) | 150 (27) | 62<br>(28) | 109 (26) | 36 (28) |
| Q4 | 127 (22) | 26 (16) | 98 (17) | 31(17) | 102<br>(20) | 39 (17) | 99 (18) | 30<br>(13) | 71 (17) | 15 (12) |
| Q5 (Highest) | 138 (24) | 26 (16) | 128 (23) | 27 (15) | 139<br>(27) | 45 (20) | 120 (22) | 17<br>(7.6) | 61 (14) | 24 (19) |

a=Ethiopia <23 deliveries/month, Kenya <20 deliveries/month and Nigeria <50 births/month; b=Ethiopia 23 to 84 deliveries/month, Kenya 20-60 deliveries/month and Nigeria 50-179 births/month; c=Ethiopia >84 deliveries/month, Kenya >60 deliveries/month and Nigeria >179 births/month; d=basic emergency obstetric and newborn care;[34] e=comprehensive emergency obstetric and newborn care;[34] f=2 individuals in Kano intervention area and one in Nasarawa control area had not specified their marital status; g=includes separated and divorced; h=missing data on religion for one individual in Machakos intervention area

### Coverage

In Ethiopia, after 12 months of the intervention, the proportion of births meeting the primary outcome was 87% (522/602) in intervention facilities and 0% in control facilities (Table 3) (X^2^=6.35, p=0.012); in Machakos it was 35% (186/526) in intervention facilities and 13% (29/224) in control facilities (X^2^=1.11, p=0.29); in Murang’a 38% (204/541) in intervention facilities and 10% (24/233) in control facilities (X^2^=1.24, p=0.27); in Kano 88% (538/611) in intervention facilities and 13% (32/251) in control facilities (X^2^=447, p<0.001); and in Nasarawa 91% (509/558) in intervention facilities and 25% (29/117) in control facilities (X^2^=260 x, p<0.001).

**Table 3.** Birth companion coverage at baseline and endline intervention and control facilities in Ethiopia, Kenya and Nigeria.

| Country | Setting | Data collection period | Study arm | Number of participants | Had a companion: |  |  |  |
| --- | --- | --- | --- | --- | --- | --- | --- | --- |
|  |  |  |  |  | During labor | At birth | Postnatal period | Continuously |
|  |  |  |  |  | n (%) | n (%) | n (%) | n (%) |
| Ethiopia | Addis Ababa/Oromia | Baseline | Intervention | N= 578 | 527 (91) | 345 (60) | 554 (96) | 305 (53) |
|  |  |  | Control | N=159 | 13 (8) | 20 (13) | 60 (38) | 2 (1) |
|  |  | Endline | Intervention | N=602 | 575 (96) | 523 (87) | 584 (97) | 522 (87) |
|  |  |  | Control | N=118 | 57 (48) | 0 (0) | 80 (68) | 0 (0) |
| Kenya | Machakos | Baseline | Intervention | N=564 | 310 (55) | 165 (29) | 315 (56) | 161 (29) |
|  |  |  | Control | N=182 | 164 (90) | 44 (24) | 161 (89) | 41 (23) |
|  |  | Endline | Intervention | N=526 | 454 (86) | 206 (39) | 448 (85) | 186 (35) |
|  |  |  | Control | N=224 | 204 (91) | 29 (13) | 219 (98) | 29 (13) |
|  | Murang'a | Baseline | Intervention | N=517 | 77 (15) | 28 (5) | 41 (8) | 25 (5) |
|  |  |  | Control | N=230 | 46 (20) | 8 (4) | 31 (14) | 6 (3) |
|  |  | Endline | Intervention | N=541 | 389 (72) | 211 (39) | 347 (64) | 204 (38) |
|  |  |  | Control | N=233 | 200 (86) | 25 (11) | 197 (85) | 24 (10) |
| Nigeria | Kano | Baseline | Intervention | N=553 | 227 (41) | 212 (38) | 227 (41) | 211 (38) |
|  |  |  | Control | N=225 | 33 (15) | 31 (14) | 33 (15) | 31 (14) |
|  |  | Endline | Intervention | N=611 | 567 (93) | 540 (88) | 574 (94) | 538 (88) |
|  |  |  | Control | N=251 | 32 (13) | 32 (13) | 32 (13) | 32 (13) |
|  | Nasarawa | Baseline | Intervention | N=422 | 320 (76) | 262 (62) | 328 (78) | 257 (61) |
|  |  |  | Control | N=127 | 71 (56) | 64 (50) | 69 (54) | 61 (48) |
|  |  | Endline | Intervention | N=558 | 513 (92) | 513 (92) | 517 (93) | 509 (91) |
|  |  |  | Control | N=117 | 29 (25) | 29 (25) | 29 (25) | 29 (25) |

In Ethiopia a logistic regression was not possible as no women in control facilities at endline had a continuous birth companion. After adjusting for clustering and endline differences, women in intervention clusters in Machakos had increased odds of having a continuous birth companion (AOR 3.7, 95% CI= 1.2-12) compared to those in control clusters, while in Murang’a the increase was not statistically significant (AOR 5.3, 95% CI=0.6-50). After adjusting for clustering, women in intervention clusters in Kano and Nasarawa had increased odds of having a continuous birth companion (Kano AOR 3.9, 95% CI= 2.5-5.3; Nasarawa <u>AOR 3.5, 95% CI=1.2-5.7)</u> compared to those in control clusters (S1 Table).

The presence of a birth companion at different stages of labor and birth in intervention and control clusters at baseline and endline are also shown in Table 3. In intervention facilitates at endline, birth companion presence increased more during birth in Ethiopia and Kano of Nigeria, while in Machakos and Murang’a the increase was greater during labor and after birth. In Nasarawa of Nigeria the increase is comparable at all three time points.

After adjusting for clustering and differences between intervention and control sites at baseline and endline, difference-in-difference analysis showed that the percentage points increase in continuous companionship was 38 (95% CI=12-66) in Ethiopia, 9 in Machakos (95% CI= -1-20), 15 in Murang’a (95% CI= 7-26), 51 in Kano (95% CI= 9-93), and 53 in Nasarawa (95% CI=11-96) (Table 4).

**Table 4:** Difference-in-difference analysis of primary outcome by intervention and control facilities in Ethiopia, Kenya and Nigeria.

|  | Baseline |  |  | Endline |  |  | Difference<br>in<br>difference*<br>[95% CI] | Difference<br>in<br>difference**<br>[95% CI] | p-<br>value |
| --- | --- | --- | --- | --- | --- | --- | --- | --- | --- |
|  | Intervention | Control | Difference | Intervention | Control | Difference |  |  |  |
|  | %<br>[95% CI] | %<br>[95%<br>CI] |  | %<br>[95% CI] | %<br>[95%<br>CI] |  |  |  |  |
| Had continuous companion |  |  |  |  |  |  |  |  |  |
| Addis<br>Ababa/Oromia,<br>Ethiopia | 53<br>[49-57] | 1<br>[0.3-5] | 52 | 87<br>[84-89] | 0 | 87 | 35<br>[11-59] | 38<br>[12-66] | 0.009 |
| Machakos,<br>Kenya | 29<br>[25-32] | 23<br>[17-29] | 6 | 35<br>[31-40] | 13<br>[8-18] | 22 | 16<br>[-3-26] | 9<br>[-1-20] | 0.077 |
| Murang'a,<br>Kenya | 5<br>[3-7] | 3<br>[12-56] | 2 | 38<br>[34-42] | 10<br>[7-15] | 27 | 25<br>[17-47] | 15<br>[7-26] | 0.002 |
| Kano,<br>Nigeria | 38<br>[10-67] | 14<br>[11-39] | 24 | 88<br>[80-96] | 13<br>[-1-26] | 75 | 51<br>[9-93] | -- *** | 0.018 |
| Nasarawa,<br>Nigeria | 61<br>[23-98] | 48<br>[22-74] | 13 | 91<br>[84-99] | 25<br>[13-63] | 66 | 53<br>[11-96] | -- *** | 0.014 |

\*Adjusted for clustering
\*\*Adjusted for clustering, wealth quintile and education in Ethiopia, Machakos and Murang'a, Kenya; marital status in Machakos and Murang'a; and facility delivery volume and age in Machakos \*\*\*No statistically significant different sociodemographic characteristics in both states in Nigeria

Fig 2 shows the change in continuous birth companion coverage across five surveys by study site. In all settings, an increase in continuous birth companion coverage in intervention facilities occurred between baseline and first quarter surveys, except for Nasarawa where a greater increase was seen between the first and second quarterly surveys. While in Ethiopia a similar trend was seen across the other quarterly surveys, in Kenya we observed a smaller increase, followed by a decrease in coverage between the third quarter and endline surveys, particularly Machakos. In Nigeria, both states showed a similar trend of increasing birth companion coverage. However, in Kano state, there was a decrease in birth companion coverage between the second and third quarters, followed by an increase from the third quarter to the endline.

**Fig 2:**
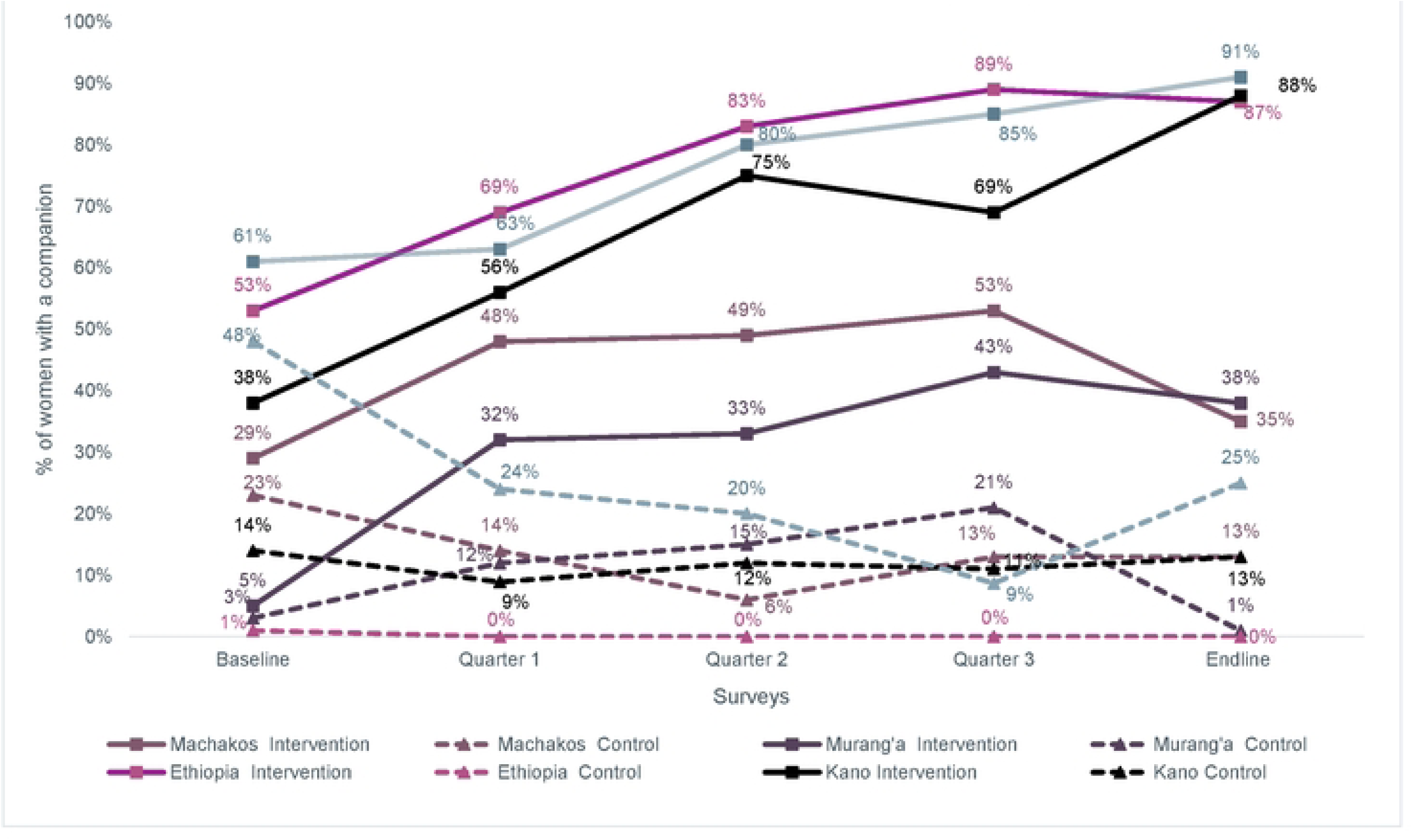
Continuous birth companion coverage tracked at five timepoints in intervention/control clusters in Ethiopia, Kenya, Nigeria.

Healthcare providers listed several challenges including that limited space and lack of privacy made it difficult to accommodate birth companions.

> *“Postnatal room the challenge we have is the space…When we have every person with a birth companion, the room becomes congested and very stuffy for the kids”* Provider_FGD_Q3_Machakos
>
> “*We have just one room used as a labor room where we admit 2-3 patients, at times if we admit up to 4 delivery women it will be hard to have those patients’ relatives at the same time all inside that labor room because the space cannot contain*” – Provider_FGD_Q3_Kano

### Types of birth companions

Some women listed more than one person as their preferred companion. In Ethiopia, most women preferred their husbands as a companion. In Machakos and Murang’a counties, most women also preferred their husbands, followed by mothers. In Nigeria, initially, most women in Nasarawa state reported preferring their husbands. However, by the endline, there was a noticeable decline in this preference with the majority of women stating they preferred their sisters, followed by their mothers (Fig 3).

**Fig 3.**
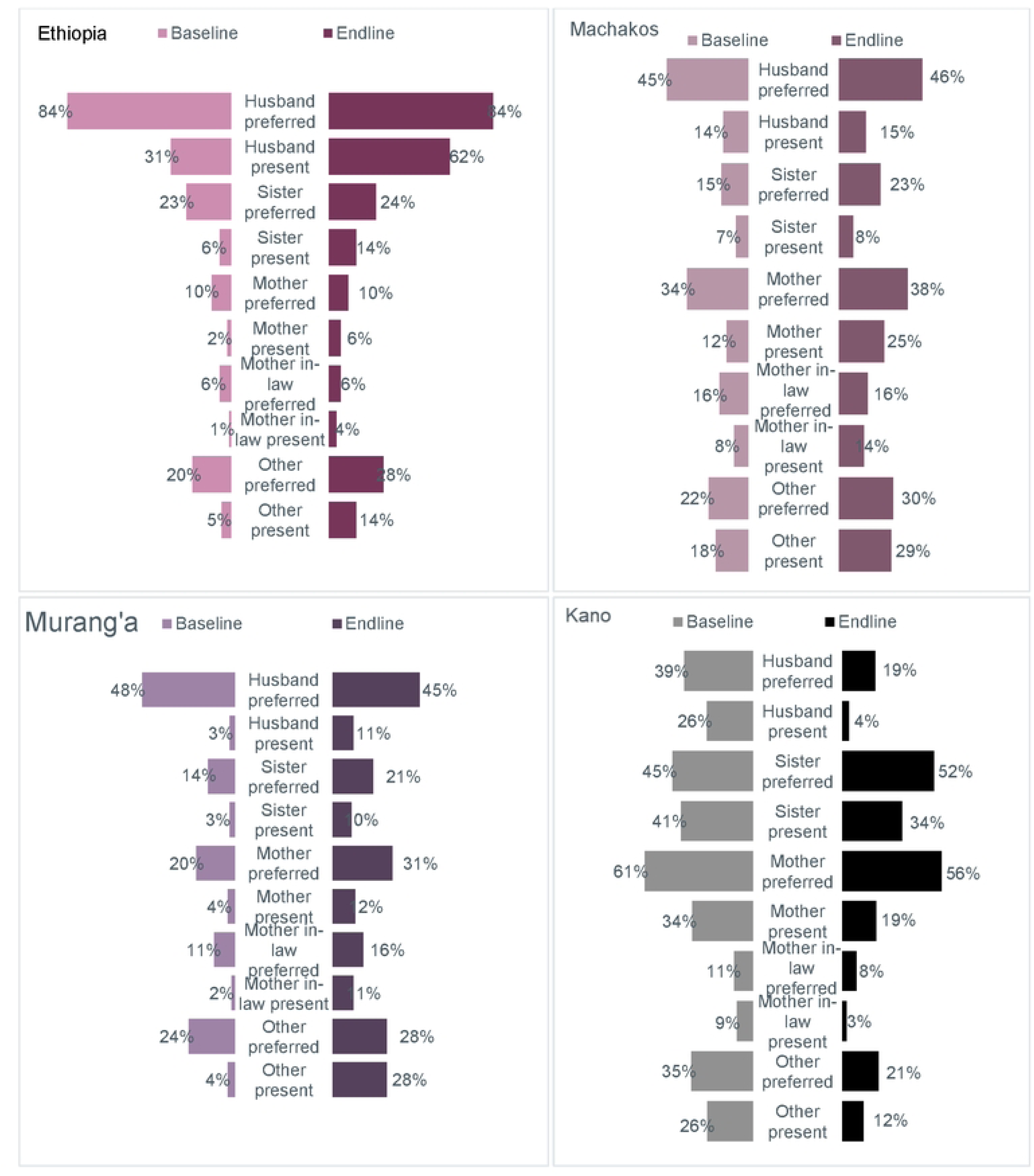
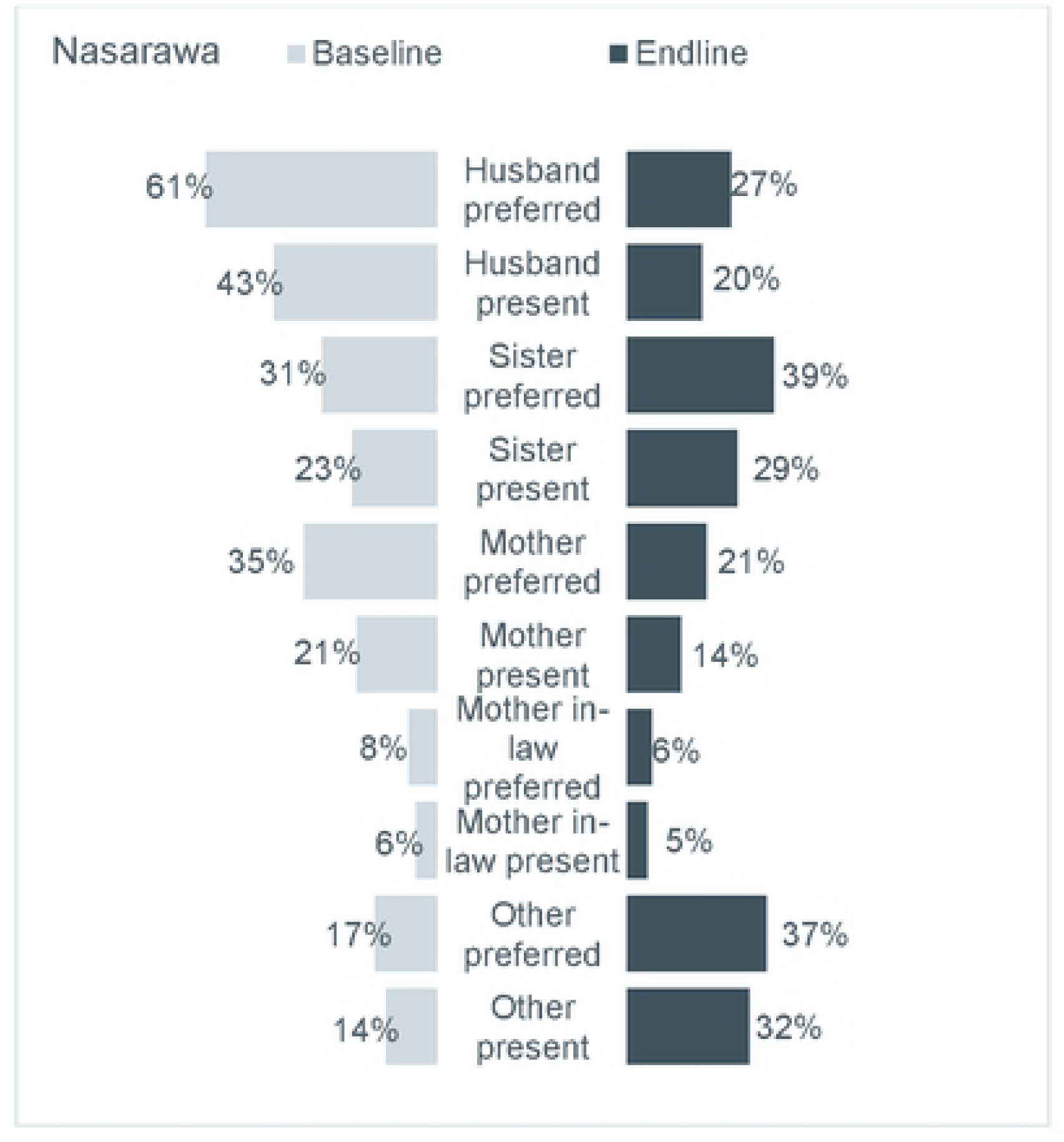
Preferred and actual birth companion continuously present during labor, birth and after birth, intervention clusters

Compared to baseline, at endline a higher proportion of women were accompanied continuously by a companion of their choice in all Ethiopia, Kenya and Kano in Nigeria, while in Nasarawa fewer women at endline were accompanied by a companion of their choice compared to baseline(Fig 3).

IDIs showed consistent findings where women reported preferring their husbands in Ethiopia and Kenya. However, in Nigeria, the interviews showed preference for sisters.

“*I chose my husband to be with me at all those times…Because there is nothing, I hide from him since he is my husband.*” Mother_IDI_Q2_Ethiopia

> *“It is him I wanted because even the mothers-in-law was around but I did not tell her, I could not even tell my friend, I wanted him to be there. Yes. Because that was his work (Laughs). He should also take that responsibility.”* Mother_IDI_Q3_Murang’a
>
> “*I prefer my elder sister from same mother and father. She is like a mother to me”*
>
> Mother_IDI_Q2_Kano

One woman, although preferring her husband, only wanted his support during specific periods.

> “*I wanted my husband, during labor and during the post-natal period, however I did not want anyone with me during delivery, so I told him to stay outside*” Mother_IDI_Endline_Ethiopia
>
> Providers also noticed that women prefer different companions at different timepoints.
>
> “*I have noticed that even if they come with their husbands, some women express a desire to have their mothers by their side during the second stage of labor [birth]… as they may feel uncomfortable with their husbands witnessing certain incidents, such as defecation, that can occur during this stage*.” Provider_FDG _Q3_Ethiopia

### Patient-centered maternity care

The 13-item PCMC scores normalized to 100 at endline are shown in S2 Table. After adjusting for differences at endline, the difference in mean PCMC score between intervention and control clusters was similar in Ethiopia and Kenya (Ethiopia: mean difference=-1.3, SE=12.4, p=0.92, Machakos: mean difference=-5.3, SE=3.1, p=0.12; and Murang’a mean difference=4.1, SE=2.7, p=0.16), while in Kano and Nasarawa there were a statistically significant mean differences (Kano: mean difference=6.6, SE=0.62, p<0.001 and Nasarawa: mean difference=12, SE=1.2, p<0.001).

The qualitative findings highlighted that most women who had a birth companion were satisfied with their experience of care regardless of their PCMC score. A woman with high PCMC score reported:

“*I didn’t think about the support I would receive from my husband before. In my previous delivery, no one was allowed to enter the room. They only asked for necessary staff to be provided but did not allow personal entry. I am happy with the change I saw.”* Mother_IDI_Q1_Ethiopia

A woman with a low PCMC score reported the following:

“*Yes, definitely. I would tell them “Even though the pain isn’t necessarily any less; you would have the strength for it if you have birth companion.” I recommend to all mothers to have someone by their side to share their experience. No one would understand them like family. Sometimes being alone is a worse feeling than the pain.”* Mother_IDI_Q3_Ethiopia

## DISCUSSION

This cluster randomized controlled trial evaluated a tailored intervention package to increase the coverage of facility births in the presence of a companion during labor, birth, and immediately after birth. We observed continuous birth companionship increased more in intervention than in control facilities. The extent of this increase varied across different sites and regions, with significant improvements observed in Ethiopia, Murang’a county, Kenya, and Kano and Nasarawa states, Nigeria. Additionally, except in Kano state, more women at the intervention facilities were accompanied by a birth companion of their choice at endline.

At baseline in Kenya, continuous birth companion coverage was similar in intervention and control facilities. At endline, compared to the control group, women in Machakos were three times more likely to have a continuous birth companion (AOR 3.4, 95% CI: 1.1-10.5) and women in Murang’a, although not statistically significant, were five times more likely to have a continuous birth companion (AOR 5.3, 95% CI: 0.6-47.0). Similar to Kenya, in Kano and Nasarawa, continuous companionship was also similar at baseline and increased by almost four folds at endline (Kano: AOR 3.9, 95% CI= 2.5-5.3; and Nasarawa: AOR 3.5, 95% CI=1.2-5.7).

At baseline in Ethiopia, despite a recognized lack of a supportive environment for the primary outcome, around half of women in the intervention facilities reported having a companion at all stages, compared to near zero coverage in control sites. This over-reporting may have stemmed from women interpreting having someone in the facility, even outside the room, as continuous companionship. To account for the difference in the primary outcome at baseline, we conducted a difference-in-difference analysis. In Ethiopia, nine in ten women had a continuous companion at endline (38-percentage points increase, 95% CI=12-66). The higher coverage of the primary outcome in Ethiopia may be attributed to various contextual factors, such as urban facilities and the presence in Ethiopia of a tertiary referral hospital with a birth companion program that supported the implementation of the intervention [35]. In Machakos, coverage increased by 9-percentage points (95% CI=-1-20) and in Murang’a by 15-percentage points (95% CI=7-26), with four in ten women in both counties having a continuous companion at endline. In Nigeria, both Kano and Nasarawa experienced a notable increase in coverage across intervention facilities. Specifically, in Kano, there was a 51-percentage points (95% CI=9-93), increase, while Nasarawa observed a 53-percentage points (95% CI=11-96), increase. <u>In</u> Nigeria, the confidence intervals widened following adjustment for clustering, attributable to intra-cluster correlation. Birth Companion coverage showed substantial variability across clusters, ranging from 0% to 100%. Additionally, unequal cluster sizes may intensify inflating the standard error, with larger facilities exerting disproportionate influence on the overall estimates. The higher coverage of birth companions in Nigeria may be attributed to the increased acceptance of birth companions in delivery rooms following the introduction of the intervention, suggesting a positive shift in local practices and attitudes toward birth companion involvement.

In Kenya, data for the quarterly surveys showed continuous birth companion coverage increased up until the third quarter survey then decreased by the endline survey, particularly in Machakos county. In Kenya, there was a healthcare provider labor strike lasting nearly eight weeks, which interrupted delivery services during the endline survey. This could have negatively affected the coverage levels at endline. For example, it is possible that the women in the study would have not received their third trimester ANC contact from the intervention facilities where they would have received information and brochures on birth companionship.

In Kenya, increasing presence of a companion during labor and after birth was feasible, but challenging during birth. This suggests that the intervention effectively addressed some but not all of the barriers to integrating birth companions into routine intrapartum care. Provider attitudes, privacy concerns, and limited delivery room space were key factors affecting companion presence during childbirth [7]. Understanding these challenges is essential for developing strategies to promote continuous birth companion presence. In Nigeria, the finding indicated intervention was able to improve companion presence at all three time points. The increased presence of birth companions during delivery by endline suggests that the intervention effectively addressed many of the challenges that had previously restricted the integration of birth companionship into routine intrapartum care. The proportion of women accompanied by a companion of choice increased across the study sites except in Kano state. The largest gap between women’s preferences and the companions they received was observed with husbands, likely due to privacy concerns, husbands’ unavailability during birth, and cultural norms around male roles during childbirth [7]. In qualitative interviews, some women expressed a preference for different companions at various stages, such as one person during labor and another during childbirth. This highlights the need for interventions to offer the option for multiple companions, including husbands, to better meet women’s diverse needs and preferences.

We observed no significant changes in PCMC scores in the Ethiopia and Kenya study settings. Including women who had a companion and those who did not in both intervention and control facilities might have diluted the association of the intervention on the experience of care. It is also possible that the presence of a companion may not have considerably affected women’s perception of their care. Companions might also not have actively engaged in the care process, underscoring the importance of orienting companions on effectively supporting women during childbirth. Additionally, the shortened PCMC tool might not have adequately captured the nuances of how companion affects their experience of care. Qualitative findings indicated that, regardless of their PCMC scores, most women who had a companion expressed satisfaction with their experience of care.

Strengths of our study included the cRCT design, which aims to minimize contamination and group differences, although total avoidance of such risks cannot be guaranteed. Inclusion of different countries and facility types provided evidence on the intervention’s feasibility in varying contexts. Our use of quarterly surveys enabled us to observe trends over time. The study design allowed for real-time programmatic adjustments. Interviews of birthing women were conducted prior to discharge, reducing recall bias. Several limitations should be noted. Despite randomization, clusters were different at baseline, which required adjustment in our analyses.

Caution should be observed in generalizing these findings. The study was limited to women who experienced vaginal births. Facilities were in an urban setting in Ethiopia, two of 47 counties in Kenya, and two of the 36 states in Nigeria which are not representative of the rest of each country. Settings without internal or external resources to reinforce implementation of the package might encounter challenges in achieving the same results.

Overall, our findings underscore the importance of context-specific strategies when implementing interventions aimed at increasing birth companion coverage. The varying degrees of birth companion coverage across the five sites highlight that while the intervention was broadly effective, it may require tailored approaches to fully realize its potential in diverse settings. Future initiatives should focus on addressing barriers to companion presence specifically during birth, involve local communities to promote birth companionship, and engage potential birth companions during pregnancy to enhance the support they can provide during childbirth.

## Contributors

DB is the principal investigator and along with AH, LN, AW, and GY was responsible for the design and development of the study. GBR, AW, JM, and AM were responsible for quantitative data analysis and GBR, SGA, MO, JM, OL and MB for qualitative data analysis. GBR and PO prepared the first draft of the manuscript with contributions from DB, FT, DB, WW, KW, DT, SS, SA, PS, SM, OK, MO, OL, ZF, MB, and AFA. All authors reviewed the manuscript and approved the final version. The roles of authors and contributors were determined according to criteria recommended by the International Committee of Medical Journal Editors.

## Data sharing

The de-identified data (URLs/accession numbers/DOIs) that support the findings of this study will be available only after acceptance of the manuscript for publication.

## Data Availability

The URLs/accession numbers/DOIs will be available only after acceptance of the manuscript for publication so that we can ensure their inclusion before publication.

## Acknowledgments

The authors express deepest gratitude to the study participants. Special thanks to field data collectors and supervisors.

## Supporting information

S1 Table Logistic regression analysis of primary outcome by intervention and control facilities at endline, Ethiopia, Kenya and Nigeria

S2 Table Linear regression results of the 13-item PCMC scale scores by endline intervention and control facilities, Ethiopia, Kenya, and Nigeria

## References

1. Trends in maternal mortality 2000 to 2020: estimates by WHO, UNICEF, UNFPA, World Bank Group and UNDESA/Population Division. [cited 13 Jun 2025]. Available: https://www.who.int/publications/i/item/9789240068759

2. A neglected tragedy: The global burden of stillbirths 2020 | UNICEF. [cited 13 Jun 2025]. Available: https://www.unicef.org/reports/neglected-tragedy-global-burden-of-stillbirths-2020

3. Hodnett ED, Gates S, Hofmeyr GJ, Sakala C. Continuous support for women during childbirth. Cochrane Database Syst Rev. 2003; CD003766. doi:10.1002/14651858.CD003766

4. WHO recommendations: intrapartum care for a positive childbirth experience. [cited 13 Jun 2025]. Available: https://www.who.int/publications/i/item/9789241550215

5. Afulani P, Kusi C, Kirumbi L, Walker D. Companionship during facility-based childbirth: results from a mixed-methods study with recently delivered women and providers in Kenya. BMC Pregnancy Childbirth. 2018;18: 150. doi:10.1186/s12884-018-1806-1

6. Bohren MA, Hofmeyr GJ, Sakala C, Fukuzawa RK, Cuthbert A. Continuous support for women during childbirth. Cochrane Database Syst Rev. 2017;7: CD003766. doi:10.1002/14651858.CD003766.pub6

7. Bohren MA, Berger BO, Munthe-Kaas H, Tunçalp Ö. Perceptions and experiences of labour companionship: a qualitative evidence synthesis. Cochrane Database Syst Rev. 2019;3: CD012449. doi:10.1002/14651858.CD012449.pub2

8. World Health Organization. WHO recommendations for augmentation of labour. Geneva: World Health Organization; 2014. Available: https://iris.who.int/handle/10665/112825

9. WHO recommendations on health promotion interventions for maternal and newborn health 2015. [cited 13 Jun 2025]. Available: https://www.who.int/publications/i/item/9789241508742

10. Utilization of birth companionship and its associated factors among laboring mothers during facilities birth in sub-Saharan Africa. Systematic review and meta-analysis - PubMed. [cited 13 Jun 2025]. Available: https://pubmed.ncbi.nlm.nih.gov/39372911/

11. Support provided by midwives to women during labour in a public hospital, Limpopo Province, South Africa: a participant observation study | BMC Pregnancy and Childbirth | Full Text. [cited 13 Jun 2025]. Available: https://bmcpregnancychildbirth.biomedcentral.com/articles/10.1186/s12884-018-1860-8

12. Perception and Experience of Post-partum Women Towards Birth Companions at a Tertiary Hospital and its Catchment Health Centers in Addis Ababa, Ethiopia | Ethiopian Journal of Reproductive Health. [cited 13 Jun 2025]. Available: https://ejrh.org/index.php/ejrh/article/view/488

13. Oluoch-Aridi J, Afulani P, Makanga C, Guzman D, Miller-Graff L. Examining person-centered maternity care in a peri-urban setting in Embakasi, Nairobi, Kenya. PLOS ONE. 2021;16: e0257542. doi:10.1371/journal.pone.0257542

14. Bohren MA, Hazfiarini A, Corona MV, Colomar M, Mucio BD, Tunçalp Ö, et al. From global recommendations to (in)action: A scoping review of the coverage of companion of choice for women during labour and birth. PLOS Global Public Health. 2023;3: e0001476. doi:10.1371/journal.pgph.0001476

15. Companion of choice during labour and childbirth for improved quality of care. [cited 13 Jun 2025]. Available: https://www.who.int/publications/i/item/WHO-SRH-20.13

16. Kenya Demographic and Health Survey - 2022 - Kenya National Bureau of Statistics. [cited 13 Jun 2025]. Available: https://www.knbs.or.ke/reports/kdhs-2022/

17. Ephi EPHI-, FMoH FM of H-, ICF. Ethiopia Mini Demographic and Health Survey 2019. 2021 [cited 13 Jun 2025]. Available: https://dhsprogram.com/publications/publication-FR363-DHS-Final-Reports.cfm

18. Health Sector Transformation in Quality.pdf. Available: http://www.dataverse.nipn.ephi.gov.et/bitstream/handle/123456789/391/Health%20Sector%20Transformation%20in%20Quality.pdf?sequence=1

19. Kenya-Health-Sector-Strategic-Plan-2018-231.pdf. Available: https://arua-ncd.org/wp-content/uploads/2022/10/Kenya-Health-Sector-Strategic-Plan-2018-231.pdf

20. PR157.pdf. Available: https://dhsprogram.com/pubs/pdf/PR157/PR157.pdf

21. Alliance WR. Respectful Maternity Care: A Nigeria-focused Health Workers’ Training Guide. Washington, DC: Futures Group, Health Policy Project. 2015.

22. Balde MD, Nasiri K, Mehrtash H, Soumah A-M, Bohren MA, Diallo BA, et al. Labour companionship and women’s experiences of mistreatment during childbirth: results from a multi-country community-based survey. BMJ Glob Health. 2020;5: e003564. doi:10.1136/bmjgh-2020-003564

23. Ige WB, Cele WB. Barriers to the provision of respectful maternity care during childbirth by midwives in South-West, Nigeria: Findings from semi-structured interviews with midwives. International Journal of Africa Nursing Sciences. 2022;17: 100449. doi:10.1016/j.ijans.2022.100449

24. Factors influencing the implementation of labour companionship: formative qualitative research in Thailand | BMJ Open. [cited 13 Jun 2025]. Available: https://bmjopen.bmj.com/content/12/5/e054946

25. Bharti J, Kumari A, Zangmo R, Mathew S, Kumar S, Sharma AK. Establishing the practice of birth companion in labour ward of a tertiary care centre in India-a quality improvement initiative. BMJ Open Qual. 2021;10: e001409. doi:10.1136/bmjoq-2021-001409

26. D B, G B, H M, F T, P O, G M, et al. A clustered randomized control trial to assess feasibility, acceptability, and impact of implementing the birth companion intervention package in Ethiopia, Kenya, and Nigeria: study protocol. BMC health services research. 2023;23. doi:10.1186/s12913-023-10082-w

27. Afulani PA, Diamond-Smith N, Golub G, Sudhinaraset M. Development of a tool to measure person-centered maternity care in developing settings: validation in a rural and urban Kenyan population. Reproductive Health. 2017;14: 118. doi:10.1186/s12978-017-0381-7

28. PASS 15 Power Analysis and Sample Size Software (2017). NCSS, LLC. Kaysville, Utah, USA, ncss.com/software/pass.

29. Afulani PA, Feeser K, Sudhinaraset M, Aborigo R, Montagu D, Chakraborty N. Toward the development of a short multi-country person-centered maternity care scale. Int J Gynaecol Obstet. 2019;146: 80–87. doi:10.1002/ijgo.12827

30. StataCorp. 2017. Stata Statistical Software: Release 15. College Station, TX: StataCorp LLC. [cited 9 Feb 2023]. Available: https://www.stata.com/support/faqs/resources/citing-software-documentation-faqs/

31. ATLAS.ti Scientific Software Development GmbH. ATLAS.ti 7.5 [Computer software]. 2015.

32. ATLAS.ti 24.0. Scientific Software Development GmbH. 2024.

33. Dedoose Version 9.0.107, cloud application for managing, analyzing, and presenting qualitative and mixed method research data (2023). Los Angeles, CA: SocioCultural Research Consultants, LLC www.dedoose.com.

34. Monitoring emergency obstetric care: a handbook. [cited 13 Jun 2025]. Available: https://www.who.int/publications/i/item/9789241547734

35. Lemma K, Bekele D, Tolu LB, Tadesse F, Kifle T, Sium AF. Birth companionship: The effect of introducing Plan-Do-Study-Act (PDSA) intervention on improving quality of care: An implementation study. Int J Gynaecol Obstet. 2025;168: 842–844. doi:10.1002/ijgo.15863

